# A Microbiome-targeting Fiber-enriched Nutritional Formula is Well Tolerated and Improves Quality of Life and Hemoglobin A1c in Type 2 Diabetes: A Double-Blind, Randomized, Placebo-Controlled Trial

**DOI:** 10.1101/2022.08.06.22278383

**Authors:** Juan P. Frias, Martin L. Lee, Ren-Hau Lai, Marc E. Washington, Christopher J. Damman

## Abstract

**OBJECTIVE:** To investigate a prebiotic fiber-enriched nutritional formula on health-related quality of life and metabolic control in type 2 diabetes.

**RESEARCH DESIGN AND METHODS:** 12-week, double-blind, placebo-controlled trial. Participants were randomized 2:1:1 to a prebiotic fiber-enriched nutritional formula (Active), a placebo fiber-absent nutritional formula (Placebo), or dietary advice alone (Diet). Primary endpoint was change in core Type 2 Diabetes Distress Assessment System (cT2-DDAS) at week 12. HbA1c change was a key secondary endpoint.

**RESULTS:** 192 participants were randomized. Mean age was 54.3 years, HbA1c 7.8%, and BMI 35.9 kg/m^2^. At week 12, cT2-DDAS (p=0.03) was reduced significantly in Active versus control arms, and HbA1c (p=0.009) was reduced significantly in Active vs Placebo arm.

**CONCLUSIONS:** A microbiome-targeting nutritional formula significantly improved cT2-DDAS and HbA1c suggesting the potential for prebiotic fiber as a complement to lifestyle and/or pharmaceutical interventions for managing type 2 diabetes.

## INTRODUCTION

Complementary approaches to traditional dietary guidance and pharmaceuticals may help curb the ever-rising epidemic of metabolic disease. Epidemiological and interventional studies point to a significant dietary fiber gap and support the benefit of high fiber foods and supplements, working in part through the microbiome, to prevent and manage metabolic disease (1–5). Rationally-designed foods and supplements that concentrate specific prebiotic fibers may provide a convenient, enjoyable, and well-tolerated complement to lifestyle and pharmaceutical approaches, improving well-being and metabolic control in persons with type 2 diabetes.

## RESEARCH DESIGN AND METHODS

This 12-week, double-blind, placebo-controlled trial randomized individuals 2:1:1 to a fiber-enriched nutritional formula (Active), a fiber-absent, iso-caloric, iso-protein nutritional formula (Placebo) or dietary advice alone (Diet). The active intervention contained a rationally-designed blend of resistant starch (RS) and oat beta-glucan (OBG) (Supplement, Formulation). This was a digital health-powered decentralized clinical trial (DCT) with enrollment representing United States’ geographic, ethnic, and racial diversity (6).

The study protocol was approved by an independent ethics committee and conducted in accordance with the principles of the Declaration of Helsinki and Good Clinical Practice guidelines. All subjects provided written informed consent before participation. The study was registered with ClinicalTrials.gov (<u>NCT05110703</u>).

Eligible subjects had self-reported type 2 diabetes for at least 90 days, HbA1c 7.5-10.5%, BMI 27-50 kg/m^2^, and were treated with diet and exercise alone or with a stable dose of antidiabetic medication (Supplement, Eligibility). Nutritional formulas were escalated over the course of 3 weeks to a maximally tolerated dose of 2 shake packets, with at least one replacing a meal, and continued for the duration of the study (Supplement, Formula Escalation). All subjects received general dietary recommendations including the <u>CDC Living With Diabetes</u> online resource (Supplement, Dietary Guidelines).

Clinical measures were obtained at baseline and week 12. The primary endpoint was the change from baseline to week 12 in the core Type 2 Diabetes Distress Assessment System (cT2-DDAS), a validated measure of diabetes distress (7). Key secondary endpoints included HbA1c, fasting plasma glucose (FPG), body weight, systolic blood pressure (SBP), and diastolic blood pressure (DBP). Other secondary endpoints included additional measures of health-related quality of life (HRQoL): World Health Organization Five Well-being Index (WHO-5) (8), Gastrointestinal Symptom Rating Scale (GSRS) (9), and an unvalidated Review of Systems Scale (ROSS) developed for this study to assess a spectrum of systemic health concerns (Supplement, HRQoL).

Sample size was determined via a difference in differences model. Assuming a difference in the mean cT2-DDAS change at 12 weeks of 0.67 standard deviations, with 80% power (and a two-sided 2.5% significance level to allow for multiple comparisons with the Active group), and a 10% dropout rate, 73 participants in the Active arm and 37 participants in the other two arms were required.

The efficacy data analysis was performed on the intent-to-treat (ITT) population, including all subjects randomized regardless of study completion. A repeated measures linear model was used that allowed for incomplete results from dropouts. Secondary endpoints such as HbA1c were evaluated similarly. Chi-square analyses were used for categorical data.

## RESULTS

The study was conducted between October 20, 2021 and May 16, 2022. Overall, 192 participants were randomized (Active, 95; Placebo, 48; Diet, 49) with additional subjects beyond sample size calculations enrolled to account for higher than anticipated dropout rate (Active, 19%; Placebo, 28%; Diet, 27%).

Demographics and clinical characteristics were comparable across arms. Overall mean (±SD) age 54.3 ± 9.7 years, duration of diabetes 8.6 ± 6.1 years, HbA1c 7.80 ± 1.62% (62 ± 13 mmol/mol), FPG 160 ± 64 mg/dL (8.89 ± 3.56 mmol/L), body weight 104.6 ± 19.5 kg, and BMI 35.9 ± 5.8 kg/m^2^. The majority of participants were female (62.0%). 62.5% were White, 19.3% Black, and 3.6% Other. 14.6% were Hispanic/Latino.

### Primary Endpoint

The cT2-DDAS change from baseline was reduced significantly in Active versus Placebo and Diet arms at week 12 (p=0.030) (Figure 1). In the Active arm, cT2-DDAS declined from a mean (±SD) of 3.1±0.9 at baseline to 2.8±0.9 at week 12. The cT2-DDAS in the Placebo arm increased from 3.0±1.0 to 3.1±0.9 and the Diet arm remained unchanged from a baseline value of 3.1±0.9. In the Active arm, there was improvement in 7 of the 8 cT2-DDAS questions (Figure 1).

**Figure 1.**
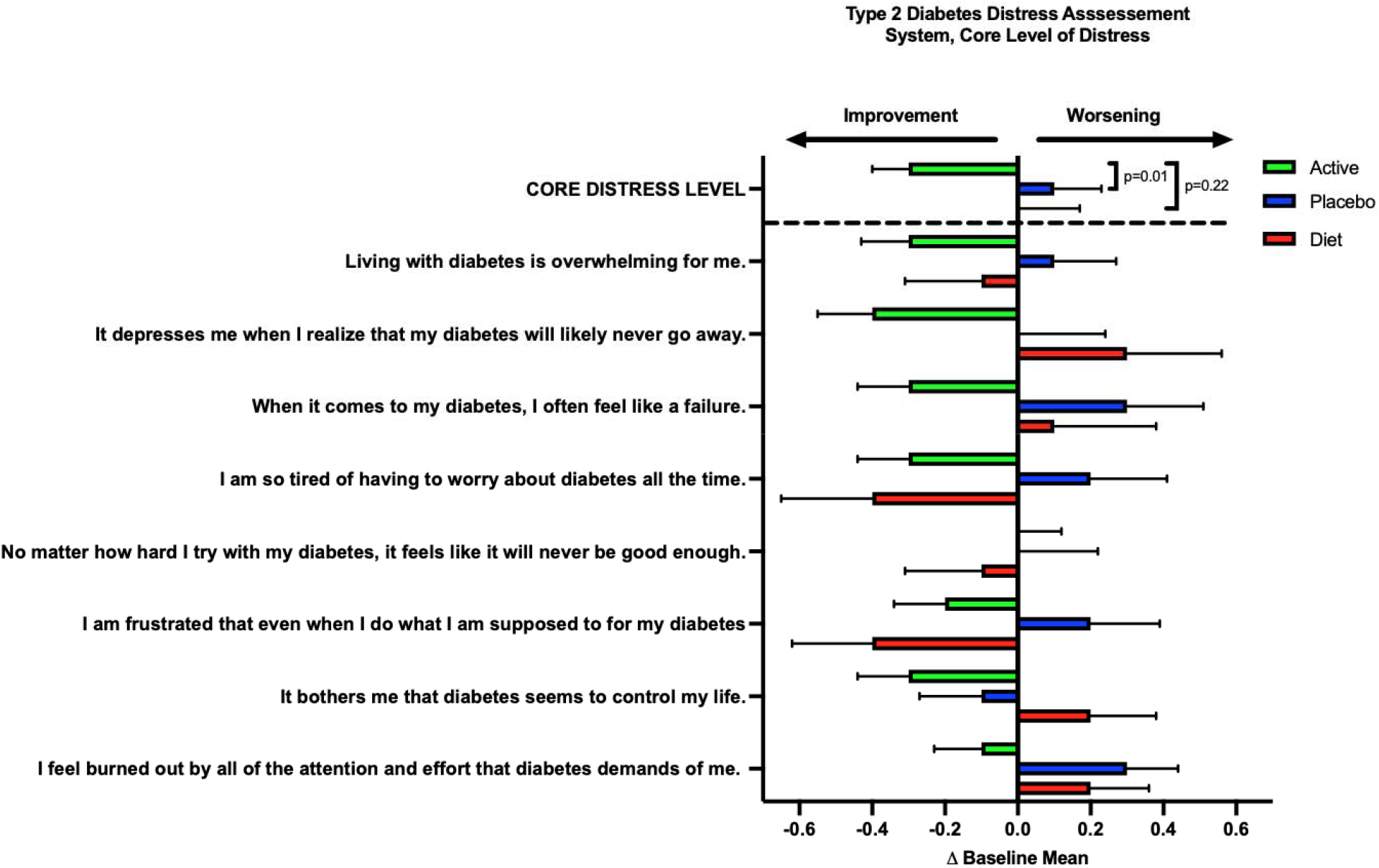
Change from baseline at week 12 in the core Type 2 Diabetes Distress Assessment System (cT2-DDAS) for the Active (green), Placebo (blue) and Diet (red) study arms. Standard errors and p-values are shown. The 3-way p-value was 0.03. A positive mean change from baseline greater than 0.25 (5% improvement) in the core distress level is considered to be clinically meaningful.

### Secondary Endpoints

At 12 weeks, the mean (±SD) change from baseline in HbA1c was -0.36±1.23%, +0.30±1.49%, and -0.17±1.26% for Active, Placebo, and Diet arms, respectively (Figure 2A). The mean difference between the change in HbA1c in the Active and Placebo arms was -0.66% (p=0.009). In participants with a baseline HbA1c of >7.0%, mean change in HbA1c at week 12 was -0.53±1.49%, +0.04±1.40%, and +0.05±1.66% for Active, Placebo, and Diet arms, respectively.

**Figure 2.**
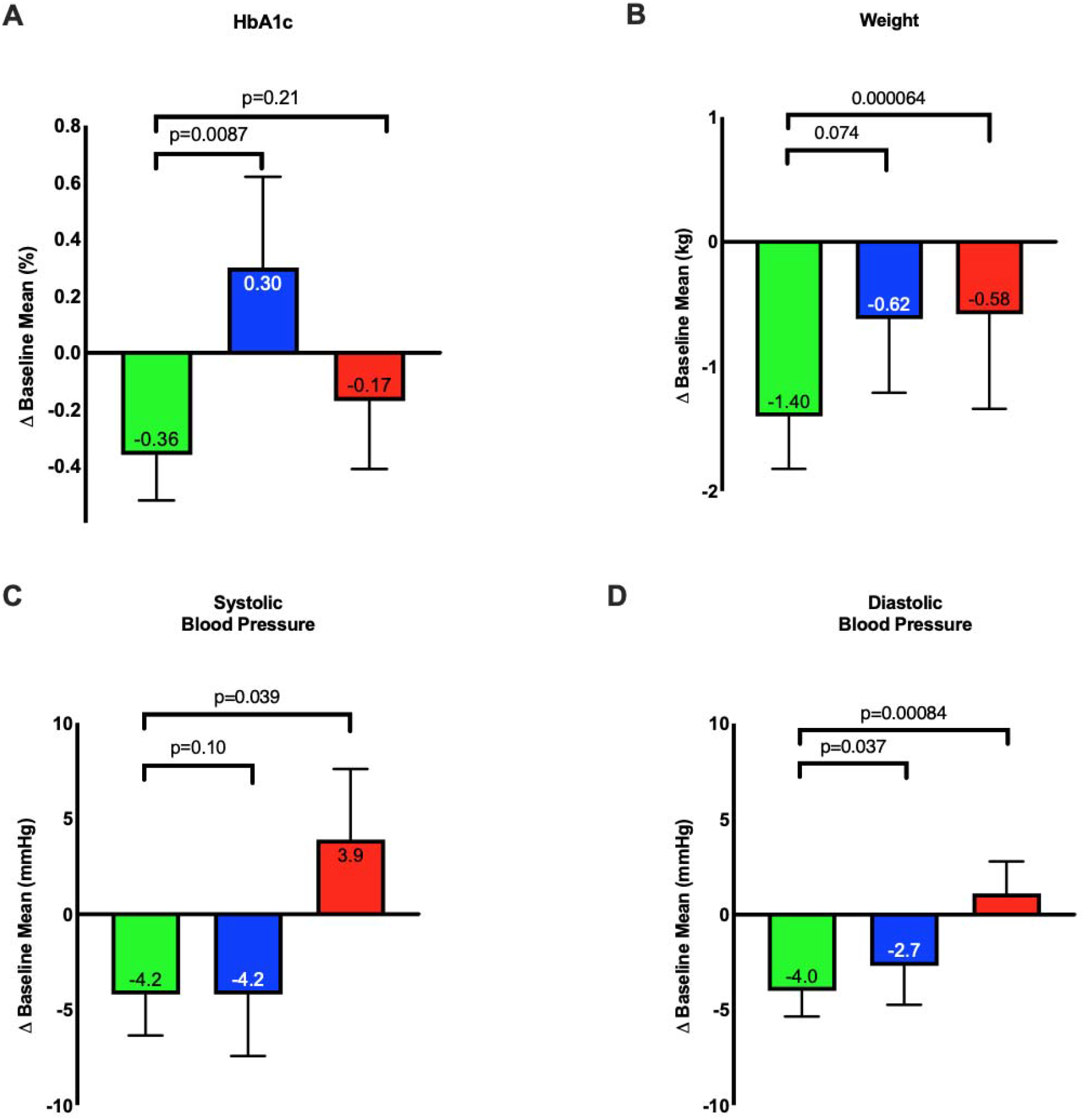
Change from baseline at week 12 in mean HbA1c (A), body weight (B), systolic blood pressure (C) and diastolic blood pressure (D) for the Active (green), Placebo (blue) and Diet (red) study arms. Standard errors and p-values are shown.

In the 3 study arms, FPG at week 12 did not change significantly from baseline and there was no difference in the change between arms. At week 12 there was a decrease in body weight in Active (1.36±3.26 kg), Placebo (0.64±2.73 kg), and Diet (0.54±3.93 kg) arms (Figure 2B). The Active arm demonstrated significantly greater weight reduction compared with Diet arm (p=0.03). Other metabolic (SBP and DBP) and HRQoL (WHO-5 and ROSS) measures improved significantly in Active arm relative to Diet arm (Figure 2C and 2D, Figures 3A and 3B; Supplement, Results Details, Figures 1 and 2).

**Figure 3.**
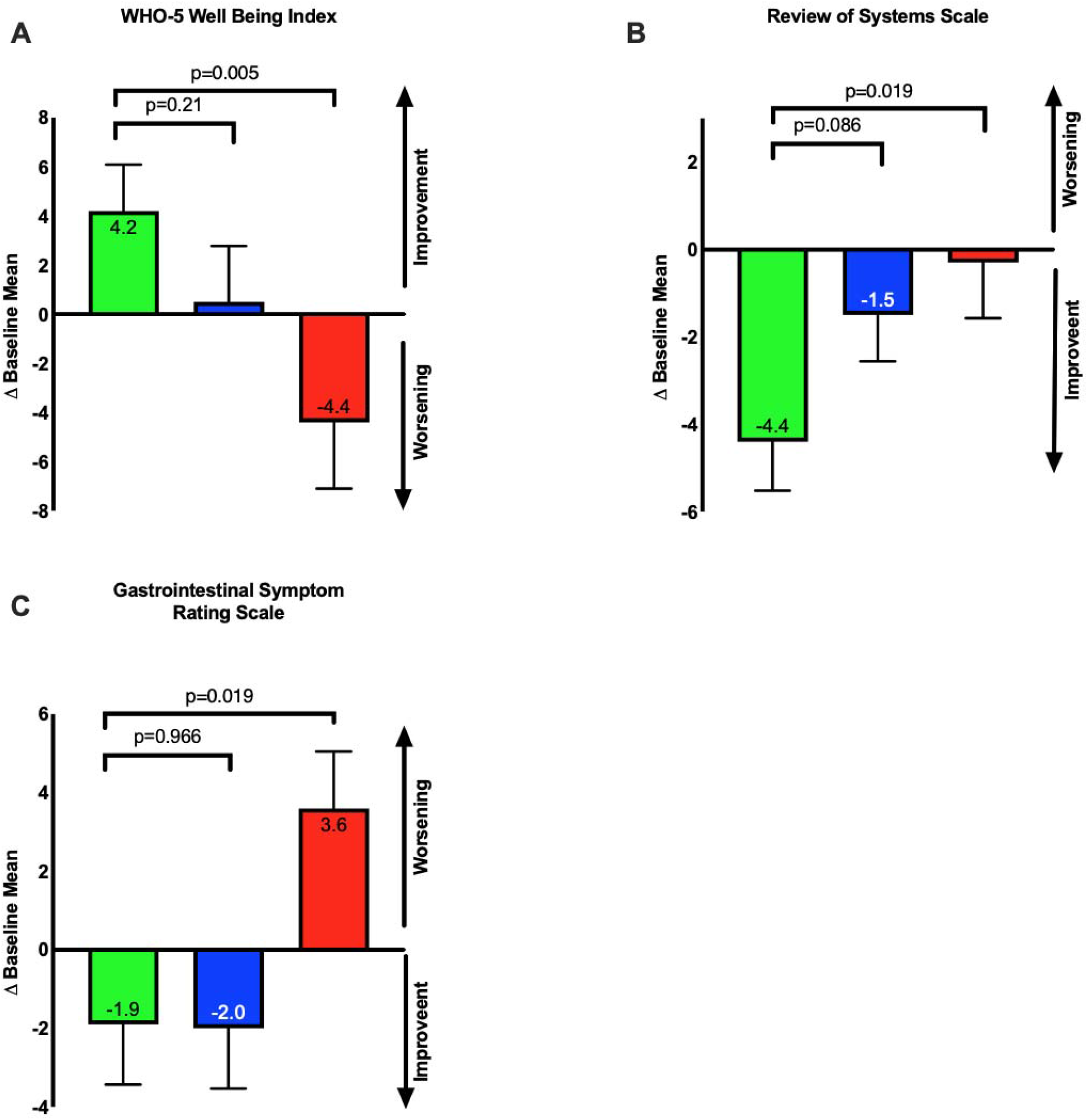
Change from baseline at week 12 in mean WHO-5 Well Being Index (A), Review of Systems Scale (B), and Gastrointestinal Rating Scale (C). Placebo (blue) and Diet (red) study arms. Standard errors and p-values are shown.

### Safety and Tolerability

There were no severe or serious adverse events and no symptomatic hypoglycemia reported. The active and placebo formulas were both well tolerated with improvement in GSRS seen in both study arms (Figure 3C, Supplement Figure 3). No subjects discontinued due to gastrointestinal intolerance. One participant in each of the Active and Placebo groups discontinued the study due to taste intolerance.

## CONCLUSIONS

In this randomized controlled exploratory trial, a microbiome-targeting fiber-enriched nutritional formula resulted in improved measures of HRQoL and metabolic health in persons with type 2 diabetes.

The primary endpoint, cT2-DDAS change from baseline at week 12, improved significantly in Active vs. Placebo and Diet arms, exceeding the minimal clinically important difference (MCID) of 0.25 recently reported by Fischer et al (10). Importantly, 7 of 8 questions contributing to the Core Level improved in the Active group. Consistent with these findings, improvements in other HRQoL questionnaires (WHO-5, ROSS) were also seen in the Active arm. These improvements may have resulted from subjects’ optimism around improved metabolic health metrics. Additionally, microbiome-derived metabolites including short chain fatty acids (SCFA’s) and neurotransmitter precursors regulate the gut-brain axis through gastrointestinal (gut permeability, inflammation, vagal stimulation) and direct central nervous system effects (blood-brain barrier permeability, neuroinflammation, and neuronal signaling) with impact on diverse neurocognitive outcomes including mood, energy, and sleep (11–13).

Consistent numeric improvements in metabolic parameters, including HbA1c, weight, and blood pressure were also seen in the Active arm. The HbA1c reduction was likely due to improvement in postprandial glucose, as no change was seen in plasma fasting glucose. Given the modest reduction in body weight (<2%), it is unlikely that this played a significant role in improving glycemic control, suggesting other mechanisms. For example, OBG related-slowing of glucose absorption due to viscosity and/or a direct effect on glucose transport via SGLT1 and GLUT1 inhibition in the small intestine (14). Improvement in metabolic parameters are also thought to result from RS and OBG’s impact on increased production of the microbiome-derived SCFA butyrate. Butyrate stimulates GLP-1 release from intestinal L-cells, enhancing insulin secretion, modulating gastric emptying and, via central mechanisms, reducing appetite (15,16). Recent epidemiological and interventional trials have suggested RS and butyrate reduce blood pressure via proposed anti-inflammatory and vagal nerve mediated mechanisms (17–19). Other differences in the Active and Placebo formulas (e.g. sweetener, fat content, and micronutrient profile) may have also impacted outcomes.

The fiber-enriched nutritional formula was well tolerated and improved GSRS. This can partly be explained by RS and OBG’s low FODMAP (fermentable oligosaccharides, disaccharides, monosaccharides and polyols) designation with limited small intestine fermentability (20). A handful of subjects experienced mild time-limited GI symptoms that were mitigated by pausing or slowing the dose escalation. There were no reported dropouts due to gastrointestinal intolerance and dropout rate was lowest in the Active arm. Most subjects discontinued for “personal reasons” including inability to comply with study procedures.

To our knowledge, this is the first double-blind, randomized controlled trial assessing a microbiome-targeted formula enriched for a blend of resistant starch and oat beta glucan on HR-QoL and metabolic outcomes in type 2 diabetes. The DCT design was an important strength, enabling geographical and racial diversity. Notably, almost 20% of participants were Black. The DCT design may also have been a limitation as it likely contributed to the higher than anticipated dropout rate. Additionally, the study was of relatively short duration and conducted during US Thanksgiving, Christmas, and Hanukkah, potentially adversely impacting metabolic outcomes. The COVID-19 Epidemic did not appear to impact outcomes. Given that inclusion criteria were self-reported, some participants had lower than anticipated HbA1c levels, but were still included in the ITT analysis. The study was not powered to assess differences in metabolic parameters.

In summary, study results suggest that a rationally-designed fiber-enriched formula containing resistant starch and oat beta-glucan is well tolerated and may serve as a complement to lifestyle and/or pharmaceutical interventions for improving quality of life in type 2 diabetes. Future studies powered for metabolic outcomes like HbA1c will be necessary to confirm these findings.

## Data Availability

All data produced in the present work are contained in the manuscript

## ACKNOWLEDGEMENTS

Acknowledgments. The authors would like to thank Citruslabs for their study coordination efforts.

## Author Contributions

C.D., J.F., M.W., M.L, and R.L. contributed to study design. J.F. provided study oversight as principal investigator. M.L. performed statistical analysis. R.L. provided guidance on use of Active and Placebo nutritional formulas. C.D. J.F., and M.L. drafted the manuscript. C.D., J.F., M.L., and M.W. edited the manuscript. C.D. is the guarantor of this work and, as such, had full access to all the data in the study and takes responsibility for the integrity of the data and the accuracy of the data analysis.

## Preprint Server

This draft manuscript has been posted on the preprint server MedRxiv.

## Prior Presentation

None.

## Online-Only Supplemental Materials

### RESEARCH DESIGN AND METHODS

#### Eligibility

##### Inclusion Criteria

Individuals are eligible to be included in the study only if they meet all of the following criteria at screening:

1. Male or female, between the age of 18 and 75 years, inclusive
2. Diagnosed with T2D for ≥90 days
3. HbA1c of 7.5 to 10.5%, inclusive
4. BMI of 27 to 50 kg/m^2^, inclusive
5. Treatment for T2D with lifestyle intervention only (for at least 90 days) or, if using antidiabetic medication(s), treated with a stable daily dose (for at least 90 days) of any of the following agents alone or in any combination: metformin (any formulation), sulfonylurea (e.g., glyburide, glipizide, glimepiride), DPP-4 inhibitor (e.g., sitagliptin, saxagliptin, linagliptin), SGLT-2 inhibitor (e.g., empagliflozin, canagliflozin, dapagliflozin, ertugliflozin), GLP-1 receptor agonists (e.g., liraglutide, semaglutide, dulaglutide)

##### Exclusion Criteria

Individuals are excluded from study enrollment if they meet any of the following criteria at screening:

1. Have type 1 diabetes or secondary forms of diabetes (e.g., secondary to cystic fibrosis)
2. Have a history of severe hypoglycemia or hyperglycemia requiring hospitalization within the prior 6 months
3. Have required insulin therapy for the treatment of T2D (with the exception of prior acute, temporary use during a hospitalization and/or for past treatment of gestational diabetes)
4. Treatment with any glucose-lowering agent(s) other than those stated in the inclusion criteria during a period of 90 days prior to screening
5. Receiving chronic oral steroid therapy (excluding those for skin, eyes, nose, or inhaled) or have received such therapy within 1 month of screening
6. Female who is pregnant, breastfeeding or intends to become pregnant during the course of the study
7. Participation in a clinical research trial within 30 days prior to screening
8. Food allergies to ingredients in the formula including but not limited to milk protein allergy
9. Ankylosing spondylitis, Crohn’s disease, and/or Celiac disease
10. Cardiovascular (CV) conditions within 2 months prior to screening: acute myocardial infarction, cerebrovascular accident (stroke), or hospitalization due to congestive heart failure (CHF)
11. Other gastrointestinal conditions which in the investigator’s opinion may jeopardize the individual’s safety or interfere with the ability to comply with the study.
12. Gastrointestinal surgeries such as those for weight loss, large bowel resection or small bowel resection which in the investigator’s opinion may jeopardize the individual’s safety or interfere with the ability to comply with the study.
13. Have a history of any other condition such as known drug, alcohol abuse, or psychiatric disorder which in the investigator’s opinion may jeopardize the individual’s safety or interfere with the ability to comply with the study.
14. Any disorder, unwillingness or inability, which in the investigator’s opinion, might jeopardize the individual’s safety or interfere with the ability to comply with the study.

#### Formulation

The fiber blend in the active nutritional formula was rationally-designed to contain multiple forms of resistant starch (RS) and oat beta glucan (OBG) representing both structural and nutritional components of plant cells that target pleiotropic mechanisms of glucose control including glucose absorption via small intestine glucose transport (SGLT1 and GLUT1 inhibition) and large intestine modulation of metabolically active microbiome metabolites. Both active and placebo formulas were in the form of a nutritional shake.

##### Dairy Vanilla Active Formula & Placebo Formula

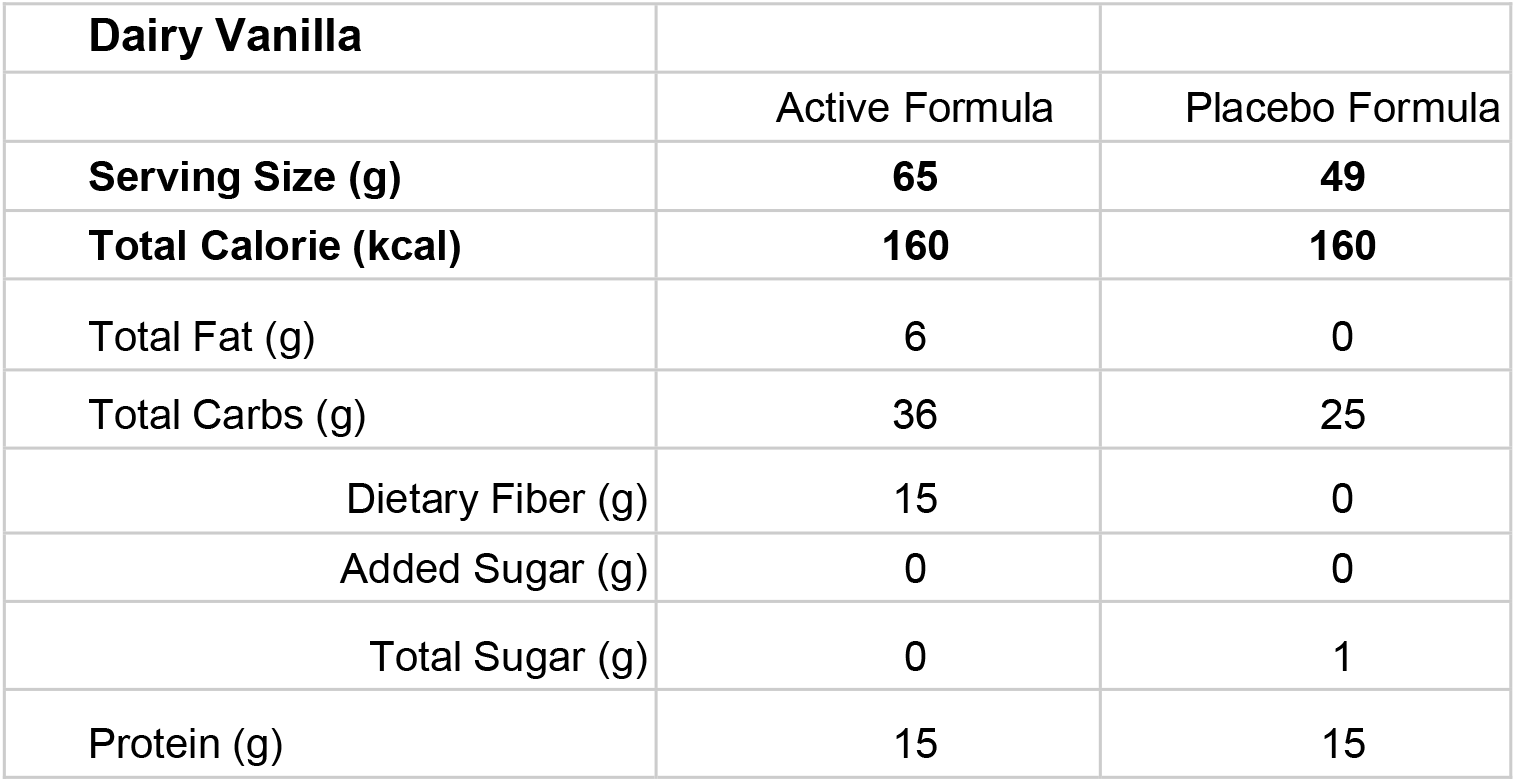

| Dairy Vanilla |  |  |
| --- | --- | --- |
|  | Active Formula | Placebo Formula |
| **Serving Size (g)** | **65** | **49** |
| **Total Calorie (kcal)** | **160** | **160** |
| Total Fat (g) | 6 | 0 |
| Total Carbs (g) | 36 | 25 |
| Dietary Fiber (g) | 15 | 0 |
| Added Sugar (g) | 0 | 0 |
| Total Sugar (g) | 0 | 1 |
| Protein (g) | 15 | 15 |

##### Active Formula Ingredient List

Prebiotic Resistant Starch Fiber Blend (Resistant Starch Blend, Soluble Vegetable Fiber, Oat Beta Glucan), Milk Protein Concentrate, Natural Sweetener Blend (Allulose, Rebaudioside M), Creamer Blend (High Oleic Sunflower Oil, Xanthan Gum, Guar Gum), Natural Vanilla Flavor, Vitamin-Mineral Blend.

##### Placebo Formula Ingredient List

Maltodextrin, Milk Protein Concentrate, Xanthan Gum, Guar Gum, Natural Vanilla Flavor, Sucralose.

##### Active Formula & Placebo Formula

| Dairy Chocolate |  |  |
| --- | --- | --- |
|  | Active Formula | Placebo Formula |
| **Serving Size (g)** | **72** | **51** |
| **Total Calorie (kcal)** | **170** | **170** |
| Total Fat (g) | 6 | 0 |
| Total Carbs (g) | 41 | 27.5 |
| Dietary Fiber (g) | 15 | 0 |
| Added Sugar (g) | 0 | 0 |
| Total Sugar (g) | 0 | 1 |
| Protein (g) | 15 | 15 |

##### Active Formula Ingredient List

Prebiotic Resistant Starch Fiber Blend (Resistant Starch Blend, Soluble Vegetable Fiber, Oat Beta Glucan), Milk Protein Concentrate, Natural Sweetener Blend (Allulose, Rebaudioside M), Creamer Blend (High Oleic Sunflower Oil, Xanthan Gum, Guar Gum), Natural Chocolate Flavor, Alkalized Cocoa Powder, Vitamin-Mineral Blend.

##### Placebo Formula Ingredient List

Maltodextrin, Milk Protein Concentrate, Xanthan Gum, Guar Gum, Natural Chocolate Flavor, Sucralose.

##### Formula Escalation

- Week 1: 0.5 shake packet of formula to replace breakfast, lunch, or dinner
- Week 2: 1 shake packet of formula to replace breakfast, lunch, or dinner
- Week 3: 1.5 shake packets of formula to replace breakfast, lunch, or dinner, and/or a snack
- Weeks 4-12: 2 shake packets of formula to replace either breakfast, lunch, or dinner, and/or snack for the remainder of the study period, effectively replacing at least one meal a day with the formula

#### Health Related Quality of Life Measures (HRQoL)

##### Type 2 Diabetes Distress Assessment System (T2-DDAS)

*Living with diabetes can be tough. Listed below are many of the stresses and worries that people with diabetes often experience. Thinking back <u>over the past month</u>, please indicate how much each of the following items were a problem for you by marking the appropriate column*.

*For example, if an item was not a problem for you over the past month, place a mark in the first column: “Not a Problem” (1). If it was a very tough problem for you, place a mark in the last column: “A Very Serious Problem” (5)*.

1. *I feel burned out by all of the attention and effort that diabetes demands of me*.
2. *It bothers me that diabetes seems to control my life*.
3. *I am frustrated that even when I do what I am supposed to for my diabetes, it doesn’t seem to make a difference*.
4. *No matter how hard I try with my diabetes, it feels like it will never be good enough*.
5. *I am so tired of having to worry about diabetes all the time*.
6. *When it comes to my diabetes, I often feel like a failure*.
7. *It depresses me when I realize that my diabetes will likely never go away*.
8. *Living with diabetes is overwhelming for me*.

##### Baseline and Endline Answers

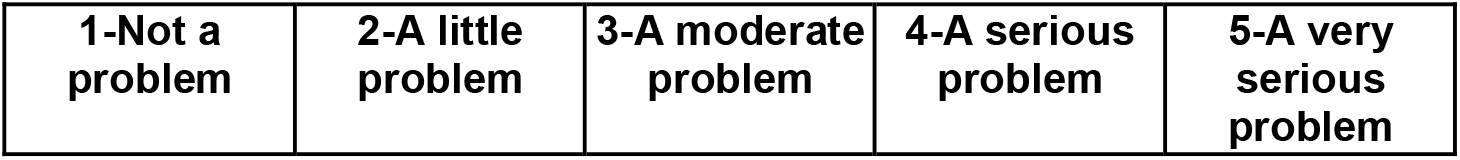

Scoring: The Core Distress Level is a single numeric score, based on the average of the eight contributing items. This score reflects the degree of, intensity, or amount of core diabetes distress reported by the respondent, with higher scores indicating greater intensity. To score, sum the scores across all eight items (1 to 5) and divide by 8 for a maximum score of 5.

Downloaded from: https://behavioraldiabetes.org/xwp/wp-content/uploads/2021/12/T2-DDAS-Core-scale-English.pdf accessed 8/4/2022.

#### WHO-5 Wellbeing Index (WHO-5)

1. *I have felt cheerful and in good spirits*
2. *I have felt calm and relaxed*
3. *I have felt active and vigorous*
4. *I woke up feeling fresh and rested*
5. *My daily life has been filled with things that interest me*

##### Baseline and Endline Answers

|  |  |  |  |  |  |
| --- | --- | --- | --- | --- | --- |
| **0-All of the time** | **1-Most of the time** | **2-More than half of the time** | **3-Less than half of the time** | **4-Some of the time** | **5-At no time** |

Scoring: WHO-5 Wellbeing Index is a sum of 5 questions scored from 0-5 and multiplied by 4 for a maximum score of 100.

#### Review of Systems Scale (ROSS)

*Which of the following health problems have bothered you in the past month?*

1. *Blood sugar levels*
2. *Weight*
3. *Digestive issues*
4. *Low energy levels*
5. *Immune health (colds/flu/viruses)*
6. *Mental health (depression/anxiety/mood)*
7. *Brain Fog*
8. *Sleep disturbances/Insomnia*
9. *Muscle or joint aches*
10. *Rashes*
11. *Allergies*
12. *Asthma*

##### Baseline Answers

Baseline Answers:
|  |  |  |  |  |
| --- | --- | --- | --- | --- |
| **1-Not at all bothered** | **2-Slightly bothered**<br>It bothers me a little, but I can go on with my life | **3-Somewhat bothered**<br>It bothers me, but it doesn't interfere with my daily life | **4-Moderately bothered**<br>It makes it hard to focus on what I'm doing | **5-Severely bothered**<br>It interferes with my daily activities |

##### Endline Answers

Endline Answers:
|  |  |  |  |  |  |
| --- | --- | --- | --- | --- | --- |
| **0-Didn't realize how much it bothered me - improved now** | **1-Not at all bothered** | **2-Slightly bothered**<br>It bothers me a little, but I can go on with my life | **3-Somewhat bothered**<br>It bothers me, but it doesn't interfere with my daily life | **4-Moderately bothered**<br>It makes it hard to focus on what I'm doing | **5-Severely bothered**<br>It interferes with my daily activities |

Score: Review of Systems is a sum score of 12 questions of concerns around different dimensions of general health (e.g. sleep, energy, and mood) on a scale of 1-5 at baseline and 0-5 at endline for a maximum score of 60.

#### Gastrointestinal Symptom Rating Scale (GSRS)

*Have you been bothered by any of the following symptoms <u>in the past week</u>*:

1. *PAIN OR DISCOMFORT IN YOUR UPPER ABDOMEN OR THE PIT OF YOUR*
2. *STOMACH in the past week*
3. *HEARTBURN in the past week?*
4. *ACID REFLUX in the past week?*
5. *HUNGER PAINS or the need to eat something in between meals in the past week?*
6. *NAUSEA (wanting to throw up or vomit) in the past week?*
7. *RUMBLING (vibration or noise of the stomach) in the past week?*
8. *BLOATED in the past week?*
9. *BURPING in the past week?*
10. *PASSING GAS OR FLATUS in the past week?*
11. *CONSTIPATION in the past week?*
12. *DIARRHEA in the past week?*
13. *LOOSE STOOLS in the past week?*
14. *HARD STOOLS in the past week?*
15. *URGENT NEED TO HAVE A BOWEL MOVEMENT in the past week?*
16. *SENSATION OF NOT COMPLETELY EMPTYING THE BOWELS in the past week?*

##### Baseline and Endline Answers

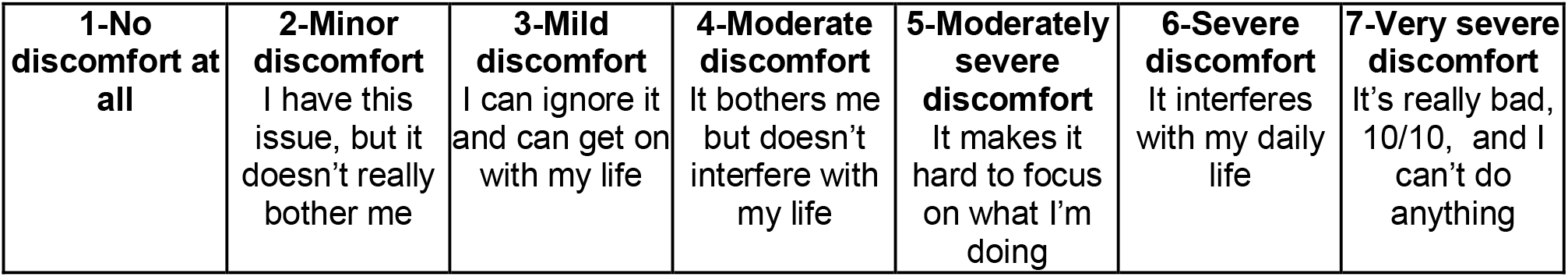

Score: Gastrointestinal Rating Scale is an average score of 15 questions of symptoms around different dimensions of gastrointestinal health on a scale of 1-7 for a maximum score of 105.

## SUPPLEMENTAL FIGURES

**Supplemental Figure 1.**
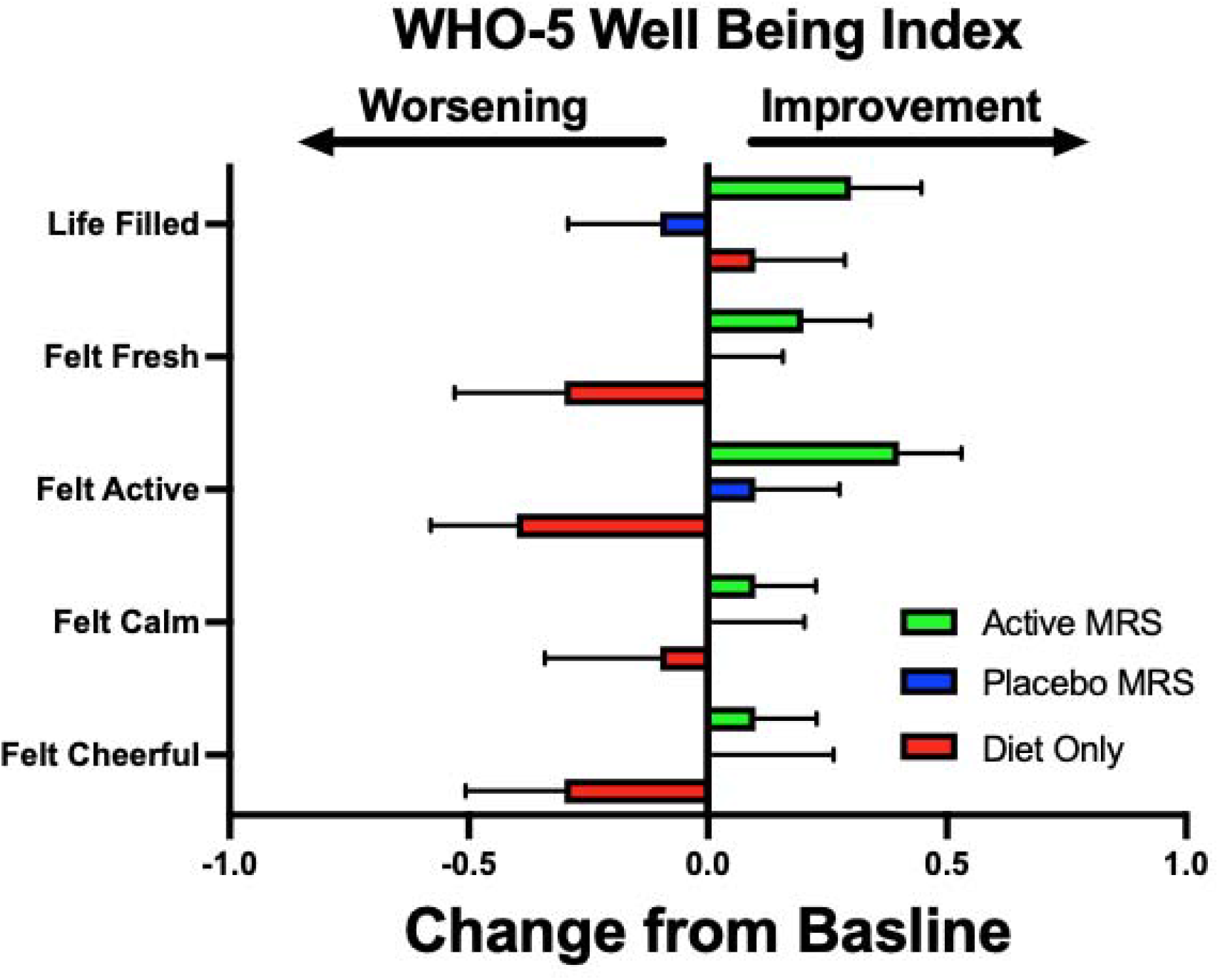
Change from baseline at week 12 in the WHO-5 Wellbeing Index (WHO-5) for the Active (green), Placebo (blue) and Diet (red) study arms. Standard errors are shown. A positive mean change from baseline of 0.5 for each question is a 10% improvement.

**Supplemental Figure 2.**
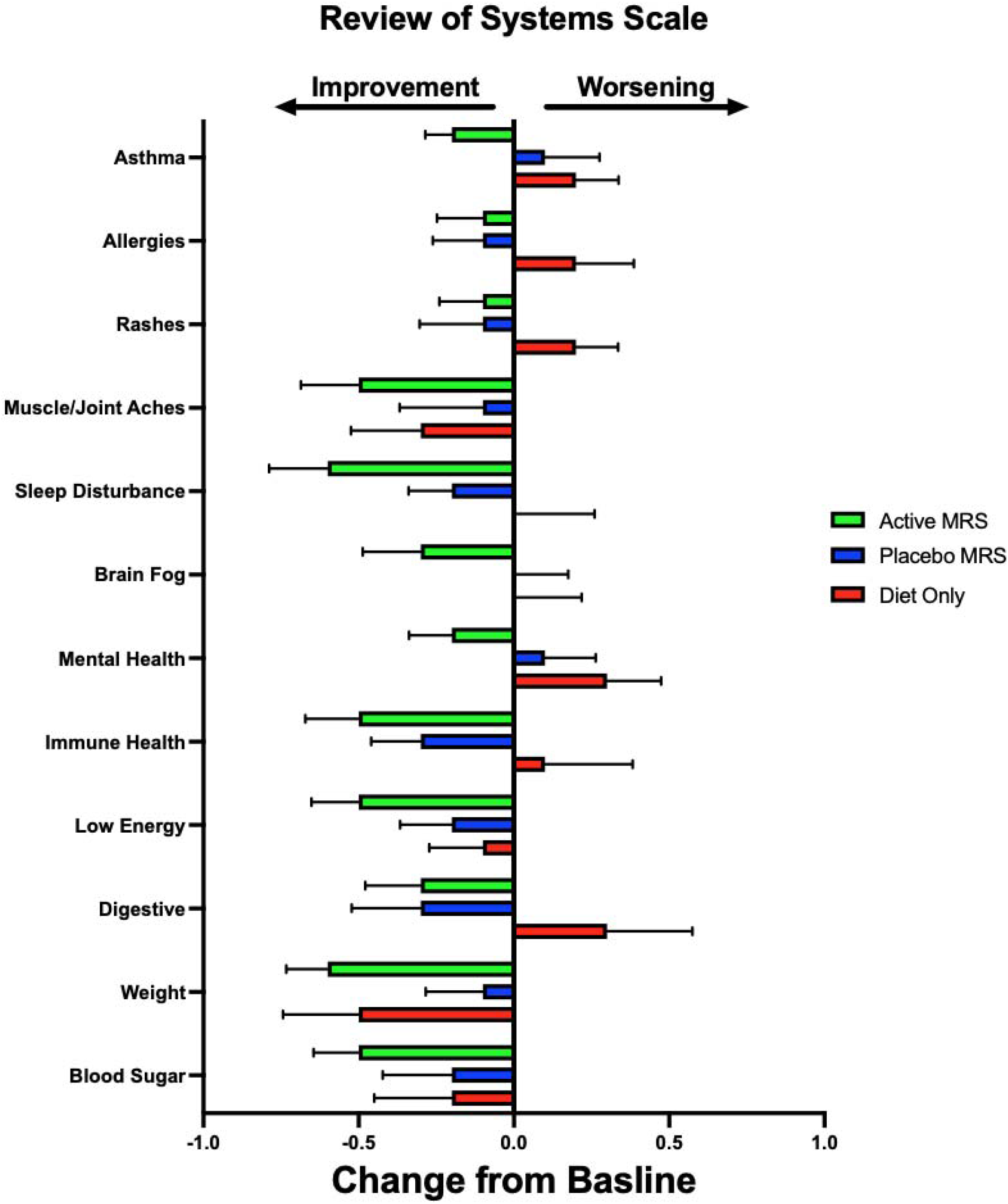
– Change from baseline at week 12 in the Review of Systems Scale (ROSS) for the Active (green), Placebo (blue) and Diet (red) study arms. Standard errors are shown. A negative mean change from baseline of 0.5 for each question is a 10% improvement.

**Supplementary Figure 3.**
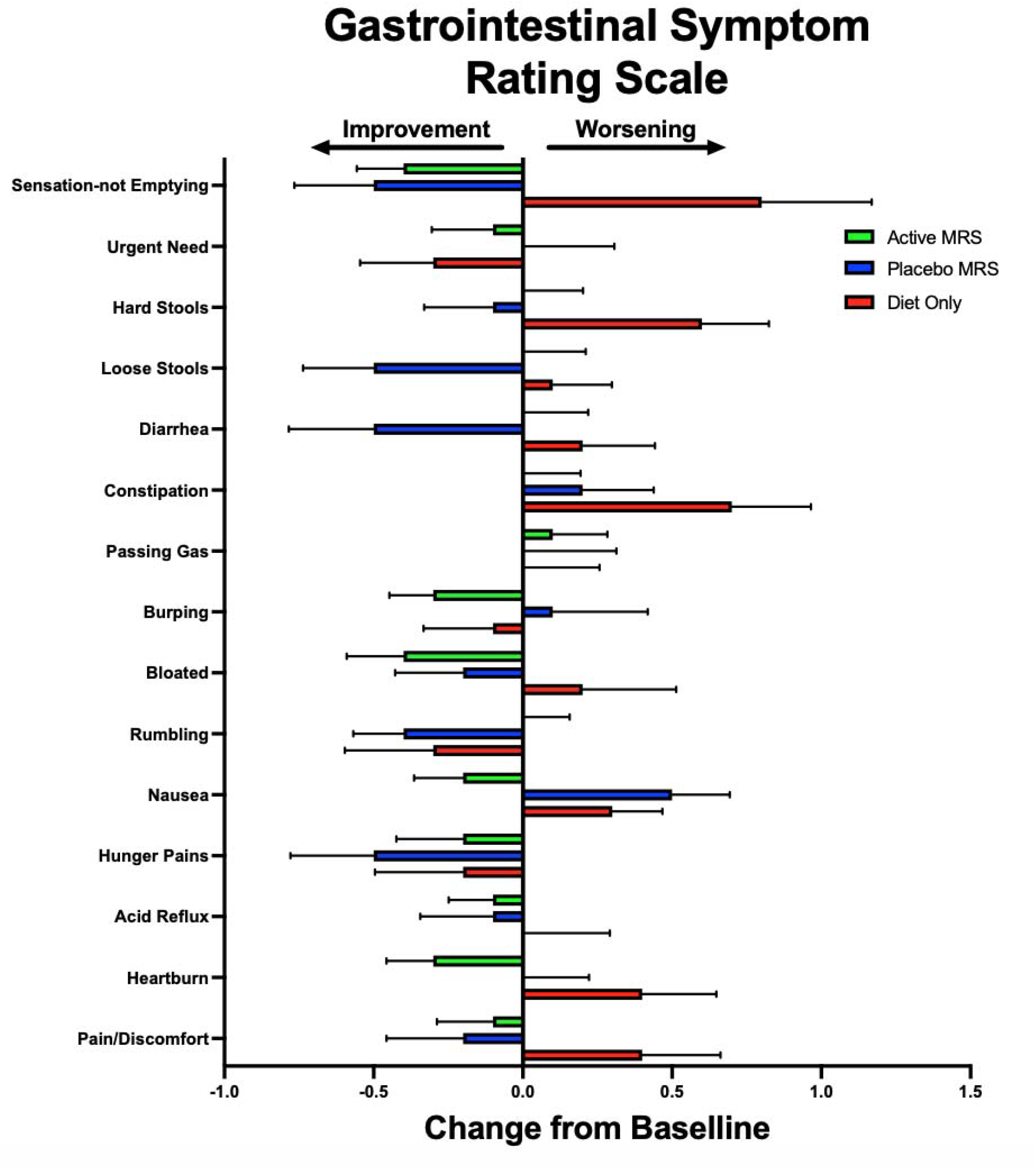
Change from baseline at week 12 in the Gastrointestinal Symptom Rating Scale (GSRS) for the Active (green), Placebo (blue) and Diet (red) study arms. Standard errors are shown. A negative mean change from baseline of 0.5 for each question is a 7% improvement.

## Dietary Guidelines - Type 2 diabetes study

As part of the study, we are providing you with these dietary guidelines that you are welcome to follow.

Healthy eating is a very important piece in managing your diabetes and the best part is that you are in control of what you eat. Many people, however, don’t really know what healthy eating actually means. Healthy eating is often associated with strict dieting that is hard to follow, restrictive, and not at all fun. Let us tell you: we are not looking for short-term changes and results (hello yo-yo effect), but a long-term change that can impact your health.

This handout should give you a better understanding of it.

Nevertheless, we recommend you talk to your certified diabetes educator (CDE), or a registered dietitian nutritionist (RDN) to help you formulate a plan that fits you and your health needs.

Whatever your taste is, watch out for nutritious food and beverages and try to avoid added sugars, sodium, alcohol, as well as saturated and trans fats.

Generally, there are 3 main types of nutrients in food: fats, proteins, and carbohydrates. Ideally, you include a portion of all these three things in each of your meals.

Here is what you want to incorporate in as many of your meals as possible:

### Carbohydrates

We don’t mean your typical white toast here, we are speaking:

- Whole-grain bread & cereals
- Oats
- Brown & Wild rice
- Quinoa
- Whole grain & enriched pasta
- Legumes (beans, peas, lentils)
- White & Sweet potatoes

### Fruits

Fruits can be an amazing alternative to dessert, as long as they are fresh, or don’t have sugar added when they’re canned. You can enjoy your fruits also frozen or dried.

### Vegetables

Did you know that most vegetables are actually superfoods? You can pick from the rainbow here: dark green, green, orange, white, yellow, purple.

### Protein & Dairy

Eat lean red meats & poultry (skinless), fish, eggs/egg whites, legumes, tofu & edamame (soy); opt for reduced-fat milk, cheese, yogurt (Greek), fortified soy/almond beverages.

### Healthy Fats & Oils

**T**he best fats come from plant sources. For cooking, opt for oils from olives, canola, walnut, peanut, soybean, sunflower; to eat, add avocado, nuts & seeds, ground flax to your diet.

#### Next steps

Especially if you’re just at the beginning of your healthy eating journey, it is important that you don’t drastically change your diet from one day to another - this is not sustainable and won’t get you any results. You have to set realistic, achievable goals and follow a diet that you enjoy and can follow long-term. Meal plans help, but we understand that sometimes it is not possible or convenient to plan everything out. Start with making healthy choices and eat, for example, oatmeal instead of pancakes for breakfast. Learn about what a serving size is and what an appropriate portion actually looks like and incorporate step by step more and more of the foods you should actually eat vs. the ones that are just pure temptations, or only represent empty calories.

The more healthy choices you make, the more your taste will change and there might even be a time when you crave fruit vs. candy; and you actually WANT to eat that greek salad vs. eating a burger with fries.

Remember that healthy eating is a journey and you and your body will need time to adjust. Don’t rush it and make long-term changes that you can follow for years to come.

The next pages are from the CDC and they discuss the following:

- Tips for eating healthy with diabetes
- Tips for being active with diabetes

For additional excellent resources on diet in diabetes please follow the link to this CDC website:

- https://www.cdc.gov/diabetes/managing/eat-well.html

